# Depression precedes problematic video streaming in adolescents: Prevalence trends and cross-lagged associations from a four-wave population-based study

**DOI:** 10.64898/2026.07.30.26359312

**Authors:** J.-O. Cloes, L. Klamert, K. Busch, K. Paschke

**Affiliations:** German Center for Addiction Research in Childhood and Adolescence (DZSKJ), University Clinic Hamburg-Eppendorf (UKE), Martinistrasse 52, D-20246 Hamburg, Germany

**Keywords:** Internet use, behavioural addictions, ICD-11, longitudinal study, epidemiology

## Abstract

**Background:** In the age of TikTok, YouTube, and Netflix, video streaming (VS) is highly popular among adolescents. Yet, risky to addiction-like viewing patterns (i.e., *problematic (P)VS*) may adversely affect well-being. Prevalence estimates based on established criteria and etiological understanding of this phenomenon remain scarce. It is associated with depression, a major issue within the youth mental health crisis. However, causality remains unclear. This study investigated prevalence trends of adolescent PVS and its temporal relationship with depression.

**Methods:** Population-based data were drawn from four annual waves (2022-2025) of a representative online survey among 3,477 German adolescents (aged 10-17 years). Weighted annual PVS prevalence estimates were calculated based on standardized measures applying ICD-11 criteria of behavioural addictions distinguishing pathological from hazardous behavioural patterns. A cross-lagged panel analysis examined the reciprocal relationship between PVS and depression over four years.

**Results:** Prevalence of *pathological VS* ranged between 2 to 4% across waves. *Hazardous VS* prevalence was 13-14% from 2022 to 2024, before increasing to 25% in 2025. Up to 81% of adolescents with *pathological VS* (21% with *hazardous VS*) showed clinically relevant symptoms of depression, versus 8-9% of non-affected adolescents. Depression significantly predicted PVS in two of three lags (β_w1-w2_=0.233, β_w2-w3_=0.155), but not vice versa.

**Conclusions:** Prevalence rates and their divergent associations with depression support distinguishing *hazardous* from *pathological VS* and underline the clinical relevance of PVS. Depression preceded PVS, pointing to the role of maladaptive coping. This has direct implications for effective intervention measures. Future research should clarify the mechanisms underlying this relationship.

## Introduction

Viewing online videos on digital platforms like Netflix, YouTube or TikTok – i.e., video streaming (VS) – is highly popular among adolescents [1]. This age group is especially susceptible to developing problematic online behaviours due to neurodevelopmental imbalances [2,3]. In 2021, approximately 11% of German 10- to 17-year-olds were affected by *problematic video streaming* (PVS, [4]), which is comparable to international prevalence rates of other forms of problematic internet use (PIU), such as problematic gaming (12%) and problematic social media use (11%) [5].

PVS seems to be of clinical relevance, as it may interfere with developmental needs in adolescence, potentially contributing to excessive usage patterns [6], and mental health impairment [7]. While the COVID-19 pandemic likely influenced PVS patterns during this time [8], updated prevalence data since 2021 or longitudinal trajectories of PVS among adolescents are currently missing, Thus, further research is warranted [9].

Additionally, operationalization of PVS is currently inconsistent [7], which increases the risk of overpathologization. However, standardized criteria for *hazardous gaming* and *gaming disorder* exist in the International Classification of Diseases (ICD-11; [10]). Consequently, these criteria have been adapted for other forms of PIU, e.g., the pathological use of social media, online pornography, and online shopping platforms [11]. Similarly, ICD-11 criteria have been applied to PVS developing instruments to assess both *hazardous* and *pathological VS* [4,12]. Research on potential risk factors of adolescent PVS is limited. Digital media use has been identified as a potential catalyst for outcomes of the global youth mental health crisis, such as rising depression rates, although the evidence is largely correlational [13]. Moreover, depression has been consistently linked to specific PVS behaviours, such as binge watching [14,15], or “addictive TikTok use” [16], as well as other forms of PIU [17–19]. In a recent cross-sectional study, depression levels were moderate for adolescents with PVS compared to healthy controls [4]; however, the causality of this relationship remained unclear due to the study design.

PVS may initially develop as a maladaptive coping strategy for depression [20,21]. Consequently, PVS might be maintained by an addicted behavioural pattern. According to the *Interaction of Person-Affect-Cognition-Execution (I-PACE) model* of internet use disorders, addictive forms of internet use develop in a dynamic interaction of predisposing variables, psychopathology (e.g., depression, social anxiety, ADHD), affective and cognitive reactions (i.e., a pleasant or aroused feeling when viewing episodes of a TV series or short videos), impairments in executive functioning (including inhibitory control), and decreased sensitivity to punishment [22,23]. While most findings on potential risk factors of PIU are cross-sectional, some longitudinal studies suggest depression to precede PIU [24–26]. However, different forms of PIU may be connected to different pathways and the true nature of this relationship remains complex and bidirectional, warranting further research.

To address these research gaps, the present study pursued two primary objectives: (1) to provide up-to-date prevalence estimates of PVS among adolescents based on standardized ICD-11 criteria and distinguishing hazardous and pathological patterns (as for *hazardous gaming and gaming disorder*) across four consecutive years (2022–2025) using representative samples of 10- to 17-year-old adolescents in Germany, and (2) to examine the temporal relationship between PVS and depression using longitudinal data over four years.

## Methods

### Participants & Procedure

Data were derived from a large population-based longitudinal online survey on digital media use and mental health in German families (i.e., adolescents and a respective parent) conducted by the German *forsa* Institute for Social Research and Statistical Analysis (for further sample descriptions, see [27,28]). For this study, data from four waves were used: June 2022 (W1), September 2023 (W2), June 2024 (W3), and September 2025 (W4). Representative samples of adolescents aged 10 to 17 years, who provided information on their weekly VS usage frequency, were collected across waves (n_w1_ = 999, n_w2_ = 1,054, n_w3_ = 999, and n_w4_ = 984) for prevalence estimations (see Supplementary Figure 1 for a flow chart).

In addition, longitudinal data from 244 adolescents participating in all four waves were used to analyse the reciprocal longitudinal relationship of adolescent PVS and depression (see Supplementary Figure 2 for a flow chart). Despite large drop-out, this subsample was comparable to the baseline sample in 2022, with no clear evidence of systematic attrition bias according to difference and equivalence testing (see Supplementary Table S1).

### Measures

*PVS* was assessed using the 10-item self-report Streaming Disorder Scale for Adolescents (STREDIS-A) adapting the ICD-11 criteria for *hazardous gaming* (ICD-11 code: QE22) and *gaming disorder* (ICD-11 code: 6C51) to assess VS patterns [4]. The scale measured cognitive-behavioural symptoms (factor 1: three items) and negative consequences (factor 2: six items). Items for both factors were scored using Likert scales ranging from strongly disagree (0) to strongly agree (4). Higher scores indicated more problematic use. An additional item assessed whether symptoms persisted for 12 months (0 = “not at all”, 1 = “only on single days”, 2 = “during longer periods”, 3 = “almost daily”). Adolescents exhibited *pathological VS* (i.e., all criteria for a disorder due to addictive behaviours were met) if they had reached the cut-off values for both factors (factor 1: cut-off > 6 points and factor 2: cut-off > 11 points) and the time criterion (cut-off > 1 point). Criteria for *hazardous VS* (i.e., a pattern of VS that increases the risk of harmful physical or mental health consequences), were fulfilled if adolescents reached the cut-off value for factor 1 but not the others. Internal consistency was excellent across waves, with Cronbach’s alpha values ranging from 0.93 to 0.94.

*Depression* was assessed using the 9-item Patient Health Questionnaire (PHQ-9; [29]). Sum scores were calculated. Cut-off values for *mild* (≥ 5 points), *moderate* (≥ 10 points), *moderately-severe* (≥ 15 points), and *severe symptoms of depression* (≥ 20 points) were used to estimate symptom severity. As commonly used, a cut-off of 10 points or higher classified adolescents with at least moderate symptom severity as having clinically relevant symptoms of depression [30]. Cronbach’s alpha values ranged from 0.88 to 0.89 across waves, indicating good internal consistency.

*Socio-demographic information* comprised age, grouped into *early* (10-13 years), *mid* (14-17 years), and *late adolescence* (18+ years) [31,32], biological sex (*female*, *male*), self-reported educational level (i.e., prospective school leaving qualification), categorised into *low* (no certificate, specialized school, or lower secondary education), *medium* (middle secondary education), and *high* (upper secondary or higher education/ university or other educational degree) based on the International Standard Classification of Education (ISCED) [33], and place of residence, separated into *rural* (below 5,000 residents) and *urban* (above 5,000 residents) according to the Federal Office for Building and Regional Planning in Germany [34].

### Statistical analysis

Analyses were conducted using R, version 4.3.2 [35]. The proportion of missing values ranged from 0-14% across variables and waves. Descriptive examination of skewness (> 2.0) and kurtosis (> 7.0) showed that all variables were within acceptable limits, supporting the assumption of normality [36]. For each wave, descriptive statistics (i.e., frequencies, means, and standard deviations) were computed using the *psych* package [37].

First, prevalence estimates for adolescent *hazardous* and *pathological VS* were calculated at each wave, using representative data of adolescents aged 10 to 17 years. Multiple imputation by chained equations (MICE) was applied to handle missing data, specifying ten imputed data sets [38]. Results were pooled using Rubin’s rules [39]. Pooled weighted prevalence estimates and corresponding confidence intervals were calculated via the *modelbased* package [40]. Population-based survey weights based on data from the Federal Statistical Office of Germany [41] were applied to adjust for adolescents’ age, biological sex, and place of residence.

Prevalence estimates were stratified by age group (*early* versus *mid adolescence*), biological sex (*female* versus *male*), and PHQ-9 (*no* to *mild symptoms of depression* versus *moderate* to *severe symptoms of depression*) using pooled estimates and contrasts. Sensitivity analyses comparing prevalence estimates based on imputed and non-imputed data showed similar results (Supplementary Table S3). Differences in stratified prevalence estimates within each wave were calculated by pairwise comparisons, while p-values were corrected for multiple comparisons using the false discovery rate (FDR) method (Table 3). Complementary, prevalence of clinically relevant symptoms of depression were estimated among PVS classifications accordingly (Table 4).

Second, a cross-lagged panel analysis (CLPA) was employed using the *lavaan* package [42] to assess the reciprocal temporal relationship between adolescent PVS and depression. For this analysis, full information maximum likelihood (FIML) was used to account for item-level missingness. Following Hamaker et al. [43], a random intercept CLPA was considered to separate within-person from between-person variance. However, persistent convergence issues precluded its use. Full metric invariance was not supported across waves (χ² (56) = 156.12, p < 0.001). Thus, single indicator latent variables were employed for STREDIS-A and PHQ-9 sum scores, justified by dominant general factors (ω_h: STREDIS-A = 0.78-0.83, PHQ-9 = 0.76-0.83), substantial explained common variance (ECV: STREDIS-A = 0.69-0.72, PHQ-9 = 0.68-0.82), and reliability (Cronbach’s alpha = 0.88-0.94). Reliability-corrected error variances were specified based on Cronbach’s alpha and observed variances. Model fit was evaluated according to established indices: comparative fit index (CFI > 0.95), Tucker-Lewis index (TLI > 0.95), root mean square error of approximation (RMSEA < 0.06), and standardized root mean square residuals (SRMR < 0.08; [44]).

The final CLPA model included autoregressive paths, indicating construct stability over time, and cross-lagged paths, representing prospective associations between constructs across waves, as well as concurrent correlations, capturing within-wave associations between constructs, estimated via maximum likelihood (ML). Age and biological sex were included as covariates to control for potential differences in STREDIS-A and PHQ-9 total scores. Effects were interpreted for cross-lagged paths (small = 0.03, moderate = 0.07, and large = 0.12), and for concurrent correlations (small = 0.12, moderate = 0.26, large = 0.48) according to Orth et al. [45].

## Results

### Sample characteristics

Weighted sociodemographic information across all four waves regarding age, biological sex, education, place of residence, average weekly VS time, PVS (STREDIS-A sum score), and depression (PHQ-9 sum score) are displayed in Table 1. Supplementary Table S2 depicts the weighted, non-imputed information.

**Table 1.** Weighted sociodemographic characteristics of the participating adolescents.

| Variables | 2022 | 2023 | 2024 | 2025 |
| --- | --- | --- | --- | --- |
|  | n = 999 | n = 1,054 | n = 999 | n = 984 |
|  | % /<br>M (SD) | % /<br>M (SD) | % /<br>M (SD) | % /<br>M (SD) |
| **Age** (in years) | 13.52 (2.24) | 13.55 (2.28) | 13.46 (2.27) | 13.51 (2.24) |
| 10-13 years | 49.93 | 49.45 | 50.08 | 49.72 |
| 14-17 years | 50.07 | 50.55 | 49.92 | 50.28 |
| **Sex** |  |  |  |  |
| Female | 48.39 | 48.64 | 48.55 | 50.30 |
| Male | 51.61 | 51.36 | 51.45 | 49.70 |
| **Education** |  |  |  |  |
| Low | 11.44 | 12.75 | 10.90 | 7.78 |
| Medium | 38.59 | 30.19 | 31.64 | 31.75 |
| High | 49.97 | 57.06 | 57.46 | 60.47 |
| **Place of residence** |  |  |  |  |
| Urban | 79.92 | 79.27 | 79.81 | 78.79 |
| Rural | 20.08 | 20.73 | 20.19 | 21.21 |
| **Average video streaming time** (in hours per week) | 9.14 (11.35) | 9.92 (11.84) | 9.58 (12.78) | 11.26 (13.14) |
| **Problematic video streaming** (STREDIS-A sum score) | 6.39 (7.74) | 6.69 (7.18) | 7.12 (7.40) | 8.27 (7.29) |
| **Depression** (PHQ-9 sum score) | 4.69 (5.19) | 4.89 (4.98) | 4.97 (4.68) | 4.90 (5.14) |

*Note.* Weighted sociodemographic information based on pooled results across multiple imputed datasets ( $m = 10$ ). Abbreviations: $n$ = sample size; % = relative frequency; $M$ = arithmetic mean; $SD$ = Standard Deviation; STREDIS-A = Streaming Disorder Scale for Adolescents; PHQ-9 = Patient Health Questionnaire 9-item version.

### Prevalence of hazardous and pathological VS

PVS prevalence estimates divided in hazardous and pathological patterns are shown in Table 2. Findings indicate that *hazardous VS* decreased slightly from 14.4% in 2022 to 12.82% in 2023, followed by an increase to 14.2% in 2024 and a peak in 2025 with 25.1%. *Pathological VS* increased from 2.6% in 2022 to 4.2% in 2023, but decreased to 3.3% in 2024 and settled at 3.8% in 2025 (see Figure 1A and 1C for a visual presentation).

**Figure 1.**
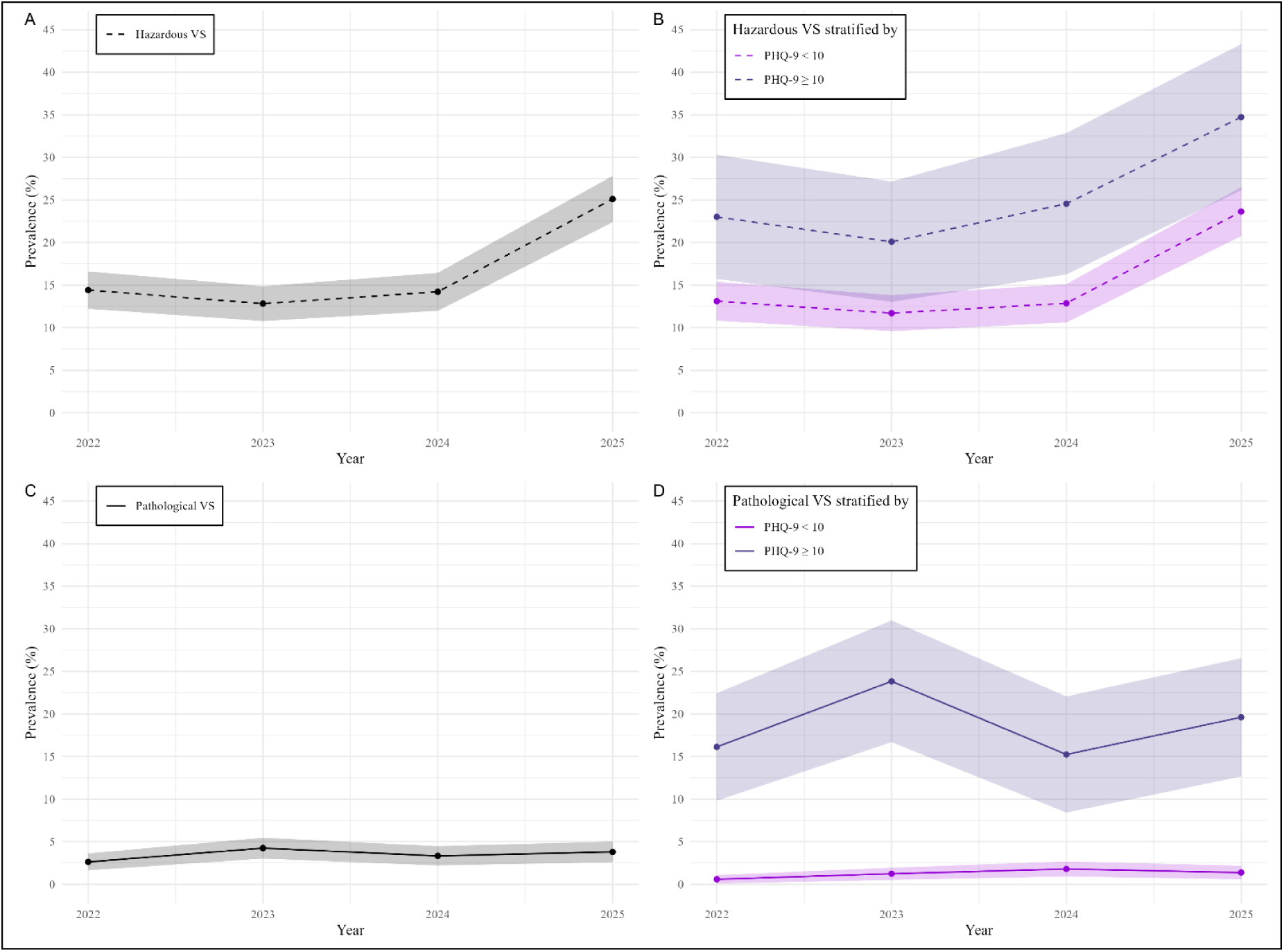
Trends of prevalence of adolescent PVS based on ICD-11 criteria between 2022 and 2025: general and stratified by depression.

**Table 2.** Weighted prevalence estimates of PVS based on ICD-11 criteria between 2022 and 2025 stratified by sex, age, and depression.

| Groups | **2022**<br>n = 999 |  | **2023**<br>n = 1,054 |  | **2024**<br>n = 999 |  | **2025**<br>n = 984 |  |
| --- | --- | --- | --- | --- | --- | --- | --- | --- |
|  | Estimate %<br>[95 % CI] |  | Estimate %<br>[95 % CI] |  | Estimate %<br>[95 % CI] |  | Estimate %<br>[95 % CI] |  |
|  | Hazardous<br>VS | Pathological<br>VS | Hazardous<br>VS | Pathological<br>VS | Hazardous<br>VS | Pathological<br>VS | Hazardous<br>VS | Pathological<br>VS |
| **Total** | 14.41<br>[12.21; 16.61] | 2.62<br>[1.63; 3.62] | 12.82<br>[10.77; 14.86] | 4.24<br>[3.01; 5.46] | 14.21<br>[11.97; 16.44] | 3.33<br>[2.20; 4.47] | 25.11<br>[22.39; 27.82] | 3.80<br>[2.59; 5.01] |
| **Sex:** |  |  |  |  |  |  |  |  |
| Female | 16.54<br>[13.19; 19.89] | 3.10<br>[1.55; 4.65] | 15.35<br>[12.18; 18.53] | 4.70<br>[2.85; 6.55] | 13.18<br>[10.00; 16.36] | 4.21<br>[2.42; 6.01] | 26.76<br>[22.84; 30.67] | 2.63<br>[1.22; 4.03] |
| Male | 12.41<br>[9.55; 15.28] | 2.17<br>[0.91; 3.44] | 10.42<br>[7.79; 13.04] | 3.80<br>[2.18; 5.41] | 15.18<br>[12.02; 18.33] | 2.51<br>[1.10; 3.91] | 23.44<br>[19.67; 27.21] | 4.99<br>[3.02; 6.97] |
| **Age:** |  |  |  |  |  |  |  |  |
| 10-13 years | 14.23<br>[11.15; 17.32] | 2.48<br>[1.11; 3.84] | 14.48<br>[11.39; 17.57] | 3.22<br>[1.69; 4.76] | 14.75<br>[11.59; 17.92] | 2.53<br>[1.13; 3.94] | 27.36<br>[23.40; 31.33] | 3.27<br>[1.68; 4.86] |
| 14-17 years | 14.59<br>[11.45; 17.73] | 2.77<br>[1.33; 4.21] | 11.19<br>[8.46; 13.92] | 5.23<br>[3.33; 7.14] | 13.66<br>[10.48; 16.84] | 4.14<br>[2.37; 5.90] | 22.88<br>[19.17; 26.58] | 4.33<br>[2.51; 6.15] |
| **Depression:** |  |  |  |  |  |  |  |  |
| No (PHQ-9 < 10) | 13.11<br>[10.8; 15.39] | 0.58<br>[0.07; 1.08] | 11.70<br>[9.58; 13.82] | 1.23<br>[0.51; 1.94] | 12.86<br>[10.61; 15.11] | 1.79<br>[0.91; 2.67] | 23.63<br>[20.75; 26.51] | 1.37<br>[0.58; 2.17] |
| Yes (PHQ-9 ≥ 10) | 23.01<br>[15.72; 30.31] | 16.13<br>[9.81; 22.44] | 20.09<br>[13.02; 27.16] | 23.84<br>[16.69; 30.99] | 24.56<br>[16.25; 32.86] | 15.24<br>[8.41; 22.06] | 34.73<br>[26.15; 43.31] | 19.61<br>[12.66; 26.56] |

*Note.* Weighted prevalence estimates are based on pooled results across multiple imputed datasets (m = 10). Results were combined using Rubin's rules. Abbreviations: PVS = Problematic Video Streaming, ICD-11 = International Classification of Diseases 11th Revision, n = sample size, % = prevalence, 95% CI = 95% Confidence Interval, VS = Video Streaming, PHQ-9 = Patient Health Questionnaire 9-item version.

On a descriptive level, female adolescents showed higher prevalence estimates for *hazardous* and *pathological VS* compared to males. Younger adolescents exhibited higher prevalence of *hazardous VS*, while the prevalence of *pathological VS* was higher in the older age group (Table 2). However, the differences between sex and age groups were partly inconsistent and non-significant across waves (see Table 3 for a full description of the comparison of *hazardous* and *pathological VS* stratified by sex and age group).

**Table 3.** Pairwise comparisons for PVS prevalence estimates stratified by sex, age, and PHQ-9 within each wave.

| Year | PVS classification | Comparisons | $\Delta$ (%) | 95% CI | z | *p*-uncorrected | *p*-corrected |
| --- | --- | --- | --- | --- | --- | --- | --- |
| 2022 | Hazardous VS | Males vs. Females | -4.13 | [-8.52; 0.27] | -1.84 | .066<sup>+</sup> | .132 |
|  |  | 14-17 years vs. 10-13 years | 0.36 | [-4.04; 4.75] | 0.16 | .875 | .875 |
| | | PHQ-9 ≥ 10 vs. PHQ-9 < 10 | 9.90 | [2.24; 17.57] | 2.53 | **.011\*** | **.033\*** |
|  | Pathological VS | Males vs. Females | -0.93 | [-0.03; 1.07] | -0.91 | .362 | .543 |
|  |  | 14-17 years vs. 10-13 years | 0.29 | [-1.69; 2.28] | 0.29 | .770 | .875 |
| | | PHQ-9 ≥ 10 vs. PHQ-9 < 10 | 15.55 | [9.22; 21.89] | 4.81 | < **.001\*\*\*** | < **.001\*\*\*** |
| 2023 | Hazardous VS | Males vs. Females | -4.93 | [-8.52; 0.27] | -2.33 | **.020\*** | .056<sup>+</sup> |
|  |  | 14-17 years vs. 10-13 years | -3.29 | [-7.46; 0.87] | -1.55 | .121 | .145 |
| | | PHQ-9 ≥ 10 vs. PHQ-9 < 10 | 8.39 | [0.92; 3.81] | 2.20 | **.028\*** | .056<sup>+</sup> |
|  | Pathological VS | Males vs. Females | -0.90 | [-2.93; 1.07] | -0.72 | .471 | .471 |
|  |  | 14-17 years vs. 10-13 years | 2.01 | [-0.45; 4.47] | 1.60 | .109 | .145 |
| | | PHQ-9 ≥ 10 vs. PHQ-9 < 10 | 22.61 | [15.42; 29.81] | 6.16 | < **.001\*\*\*** | < **.001\*\*\*** |
| 2024 | Hazardous VS | Males vs. Females | 2.00 | [-2.49; 6.49] | 0.87 | .383 | .460 |
|  |  | 14-17 years vs. 10-13 years | -1.09 | [-5.59; 3.41] | -0.48 | .634 | .634 |
| | | PHQ-9 ≥ 10 vs. PHQ-9 < 10 | 11.70 | [3.11; 20.28] | 2.67 | **.008\*\*** | **.024\*** |
|  | Pathological VS | Males vs. Females | -1.70 | [-3.99; 0.57] | -1.47 | .142 | .243 |
|  |  | 14-17 years vs. 10-13 years | 1.61 | [-0.64; 3.85] | 1.40 | .162 | .243 |
| | | PHQ-9 ≥ 10 vs. PHQ-9 < 10 | 13.45 | [6.56; 20.33] | 3.83 | < **.001\*\*\*** | **.001\*\*** |
| 2025 | Hazardous VS | Males vs. Females | -3.32 | [-8.77; -2.13] | -1.95 | .232 | .278 |
|  |  | 14-17 years vs. 10-13 years | -4.48 | [-9.93; 0.95] | -1.62 | .106 | .159 |
| | | PHQ-9 ≥ 10 vs. PHQ-9 < 10 | 11.10 | [1.92; 20.27] | 2.37 | **.018\*** | .054<sup>+</sup> |
|  | Pathological VS | Males vs. Females | 2.36 | [-0.06; 4.79] | 1.91 | .056<sup>+</sup> | .112 |
|  |  | 14-17 years vs. 10-13 years | 1.06 | [-1.35; 3.47] | 0.86 | .387 | .387 |
| | | PHQ-9 ≥ 10 vs. PHQ-9 < 10 | 18.24 | [11.25; 25.22] | 5.12 | < **.001\*\*\*** | < **.001\*\*\*** |

*Note.* Multiply imputed data. Abbreviations: PVS = Problematic Video Streaming, PHQ-9 = Patient Health Questionnaire 9-item version, $\Delta$ (%) = difference in predicted probabilities (percentage points), 95% CI = 95% Confidence Interval, z = z-statistic, *p*-uncorrected = p-value without correction, *p*-corrected = p-values corrected for multiple testing using false discovery rate (FDR) method, VS = Video Streaming, <sup>+</sup> $p < .10$ , \* $p < .05$ , \*\* $p < .01$ , \*\*\* $p < .001$ .

### Prevalence of hazardous and pathological VS, stratified by depression

Prevalence of *hazardous VS* ranged from 20.1% to 34.7% among adolescents with clinically relevant symptoms of depression (Table 2, Figure 1B). In two out of the four waves, estimates were significantly higher compared to adolescents with no or mild symptoms of depression (2022: 23.0% versus 13.1%, Δ = 9.9%; 2024: 24.6% versus 12.9%, Δ = 11.7%; see Table 3 for complete pairwise comparisons).

Prevalence of *pathological VS* among adolescents with clinically relevant symptoms of depression ranged from 15.2% to 23.8% across waves (Table 2, Figure 1D). The prevalence was significantly higher (Δ = 13.5-22.6%) compared to adolescents with no to mild symptoms, which ranged between 0.6 and 1.8% in this group (Table 3).

Conversely, the prevalence of clinically relevant symptoms of depression among adolescents with *pathological VS* ranged between 52.5 and 80.9%, compared to 18.4 to 21.0% among adolescents with *hazardous VS* and 8.4 to 9.7% among adolescents with non-problematic VS (see Table 4 for complete group comparisons across waves). All group differences between adolescents with *hazardous* and *pathological VS* (Δ = 32.6-59.9%), and adolescent with non-problematic VS compared to *hazardous VS* (Δ = 9.9-11.9%) and *pathological VS* (Δ = 44.1-71.3%) were statistically significant.

**Table 4.** Weighted prevalence estimates of depression (PHQ-9 ≥ 10) between 2022 and 2025, stratified by PVS classifications, incl. pairwise comparisons.

| Year | PVS classification | Prevalence [95% CI] | Compared to | $\Delta$ (%) | 95% CI | z | *p*-uncorrected | *p*-corrected |
| --- | --- | --- | --- | --- | --- | --- | --- | --- |
| 2022 | Pathological VS | 80.93 [65.83; 96.03] | Hazardous VS | 59.91 | [43.38; 76.43] | 7.11 | < .001*** | < .001*** |
|  |  |  | Non-problematic VS | 71.27 | [56.01; 86.53] | 9.16 | < .001*** | < .001*** |
|  | Hazardous VS | 21.02 [14.29; 27.75] | Non-problematic VS | 11.36 | [4.29; 18.44] | 3.15 | .002** | .002** |
|  | Non-problematic VS | 9.66 [7.60; 11.72] | - | - | - | - | - | - |
| 2023 | Pathological VS | 74.93 [62.14; 87.71] | Hazardous VS | 54.05 | [39.29; 68.80] | 7.18 | < .001*** | < .001*** |
|  |  |  | Non-problematic VS | 65.92 | [52.98; 78.86] | 9.98 | < .001*** | < .001*** |
|  | Hazardous VS | 20.88 [13.56; 28.20] | Non-problematic VS | 11.87 | [4.23; 19.52] | 3.05 | .002** | .002** |
|  | Non-problematic VS | 9.01 [7.02; 10.99] | - | - | - | - | - | - |
| 2024 | Pathological VS | 52.50 [35.15; 69.86] | Hazardous VS | 32.63 | [13.76; 51.50] | 3.39 | < .001*** | .001** |
|  |  |  | Non-problematic VS | 44.11 | [26.64; 61.57] | 4.95 | < .001*** | < .001*** |
|  | Hazardous VS | 19.87 [13.02; 26.73] | Non-problematic VS | 11.48 | [4.36; 18.59] | 3.16 | .002** | .002** |
|  | Non-problematic VS | 8.40 [6.48; 10.31] | - | - | - | - | - | - |
| 2025 | Pathological VS | 68.75 [53.70; 83.80] | Hazardous VS | 50.31 | [34.46; 66.16] | 6.22 | < .001*** | < .001*** |
|  |  |  | Non-problematic VS | 60.17 | [45.02; 75.31] | 7.79 | < .001*** | < .001*** |
|  | Hazardous VS | 18.44 [13.57; 23.31] | Non-problematic VS | 9.86 | [4.36; 15.35] | 3.52 | < .001*** | < .001*** |
|  | Non-problematic VS | 8.58 [6.14; 11.03] | - | - | - | - | - | - |

*Note.* Weighted prevalence estimates are based on pooled results across multiple imputed datasets ( $m = 10$ ). Results were combined using Rubin's rules. Abbreviations: PHQ-9 = Patient Health Questionnaire 9-item version, PVS = Problematic Video Streaming, 95% CI = 95% Confidence Interval, $\Delta$ (%) = difference in predicted probabilities (percentage points), z = z-statistic, *p*-uncorrected = p-value without correction, *p*-corrected = p-values corrected for multiple testing using false discovery rate (FDR) method, VS = Video Streaming. + $p < .10$ , \* $p < .05$ , \*\* $p < .01$ , \*\*\* $p < .001$ .

### Reciprocal relationship of adolescent PVS and depression

The CLPA model demonstrated excellent fit (CFI = 0.991, TLI = 0.987, RMSEA = 0.054, SRMR = 0.048, Figure 2). Significant and large autoregressive effects were observed for both PVS (β_w1-w2_ = 0.371, β_w2-w3_ = 0.407, β_w3-w4_ = 0.374) and depression (β_w1-w2_ = 0.717, β_w2-w3_ = 0.593, β_w3-w4_ = 0.501) indicating concept stability (Table 5). Concurrent correlations between PVS and depression were moderate to large and highly significant at each wave (r_w1_ = 0.515; r_w2_ = 0.359, r_w3_ = 0.407, r_w4_ = 0.521), demonstrating a stable co-occurrence of the two construct. The analysis revealed large cross-lagged effects from depression to PVS (β_w1-w2_ = 0.233, β_w2-w3_ = 0.155, β_w3-w4_ = 0.142), while cross-lagged effects from PVS to depression were small and not significant (β_w1-w2_ = 0.043, β_w2-w3_ = 0.036, β_w3-w4_ = 0.041). Regarding covariates, age and sex showed mostly non-significant effects, except for a large negative effect of age on PVS in 2024 (β = −0.182), a large positive effect of age on depression in 2023 (β = 0.127) and a large negative effect of female sex on depression in 2022 (β = −0.140)

**Figure 2.**
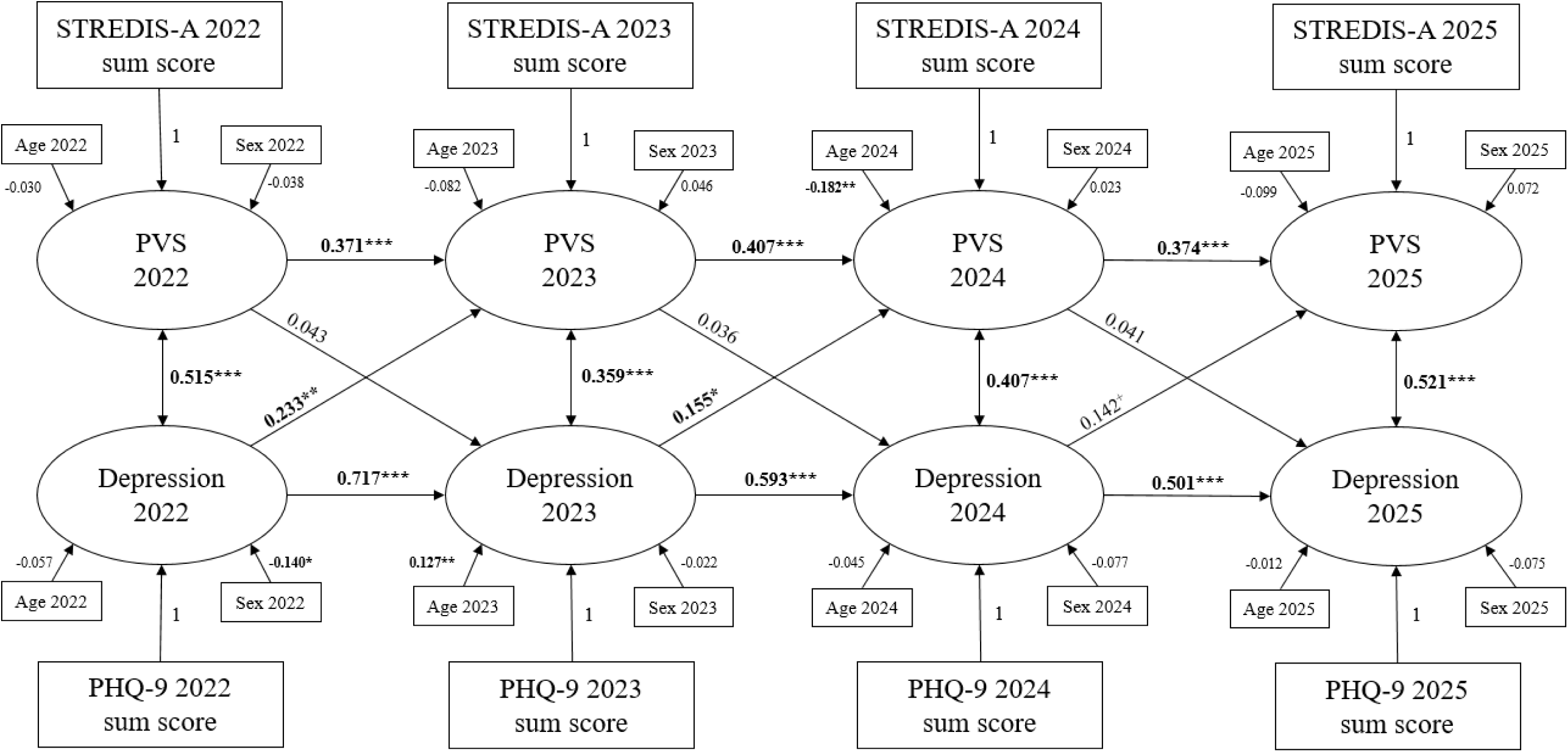
Cross-lagged panel model of the temporal relationship between adolescent PVS and depression from 2022 to 2025 in Germany (n = 244).

**Table 5.** Results of the cross-lagged panel analysis of the relationship between adolescent PVS and depression from 2022 to 2025 (n = 244).

| Path | $\beta$ | Lower 95% CI | Upper 95% CI | SE | *p* |
| --- | --- | --- | --- | --- | --- |
| **Autoregressive effects:** |  |  |  |  |  |
| PVS 2022 → PVS 2023 | 0.371 | 0.237 | 0.522 | 0.073 | < .001*** |
| PVS 2023 → PVS 2024 | 0.407 | 0.279 | 0.574 | 0.075 | < .001*** |
| PVS 2024 → PVS 2025 | 0.374 | 0.215 | 0.487 | 0.069 | < .001*** |
| Depression 2022 → Depression 2023 | 0.717 | 0.622 | 0.904 | 0.072 | < .001*** |
| Depression 2023 → Depression 2024 | 0.593 | 0.487 | 0.796 | 0.079 | < .001*** |
| Depression 2024 → Depression 2025 | 0.501 | 0.350 | 0.634 | 0.073 | < .001*** |
| **Cross-lagged effects:** |  |  |  |  |  |
| Depression 2022 → PVS 2023 | 0.233 | 0.102 | 0.447 | 0.088 | .002** |
| Depression 2023 → PVS 2024 | 0.155 | 0.012 | 0.349 | 0.086 | .036* |
| Depression 2024 → PVS 2025 | 0.142 | -0.003 | 0.290 | 0.075 | .055 |
| PVS 2022 → Depression 2023 | 0.043 | -0.077 | 0.155 | 0.059 | .506 |
| PVS 2023 → Depression 2024 | 0.036 | -0.100 | 0.170 | 0.069 | .610 |
| PVS 2024 → Depression 2025 | 0.041 | -0.094 | 0.170 | 0.067 | .573 |
| **Covariate effects:** |  |  |  |  |  |
| Age 2022 → PVS 2022 | -0.030 | -0.031 | 0.019 | 0.019 | .651 |
| Sex 2022 → PVS 2022 | -0.038 | -0.174 | 0.094 | 0.094 | .562 |
| Age 2023 → PVS 2023 | -0.082 | -0.038 | 0.006 | 0.006 | .152 |
| Sex 2023 → PVS 2023 | 0.046 | -0.072 | 0.168 | 0.168 | .431 |
| Age 2024 → PVS 2024 | -0.182 | -0.060 | -0.014 | -0.014 | .002** |
| Sex 2024 → PVS 2024 | 0.023 | -0.100 | 0.150 | 0.150 | .692 |
| Age 2025 → PVS 2025 | -0.099 | -0.040 | 0.004 | 0.004 | .101 |
| Sex 2025 → PVS 2025 | 0.072 | -0.047 | 0.197 | 0.197 | .227 |
| Age 2022 → Depression 2022 | -0.057 | -0.032 | 0.013 | 0.013 | .407 |
| Sex 2022 → Depression 2022 | -0.140 | -0.245 | -0.005 | -0.005 | .041* |
| Age 2023 → Depression 2023 | 0.127 | 0.005 | 0.040 | 0.040 | .014* |
| Sex 2023 → Depression 2023 | -0.022 | -0.118 | 0.076 | 0.076 | .673 |
| Age 2024 → Depression 2024 | -0.045 | -0.030 | 0.013 | 0.013 | .433 |
| Sex 2024 → Depression 2024 | -0.077 | -0.194 | 0.035 | 0.035 | .174 |
| Age 2025 → Depression 2025 | -0.012 | -0.023 | 0.019 | 0.019 | .840 |
| Sex 2025 → Depression 2025 | -0.075 | -0.194 | 0.042 | 0.042 | .208 |

| Path | r | Lower 95% CI | Upper 95% CI | SE | p |
| --- | --- | --- | --- | --- | --- |
| **Concurrent correlations:** |  |  |  |  |  |
| Depression 2022 ~~ PVS 2022 | 0.515 | 0.385 | 0.645 | 0.017 | < .001*** |
| Depression 2023 ~~ PVS 2023 | 0.359 | 0.194 | 0.524 | 0.012 | < .001*** |
| Depression 2024 ~~ PVS 2024 | 0.407 | 0.255 | 0.559 | 0.014 | < .001*** |
| Depression 2025 ~~ PVS 2025 | 0.521 | 0.368 | 0.674 | 0.015 | < .001*** |

*Note.* Model fit: CFI = 0.991, TLI = 0.987, RMSEA = 0.054, SRMR = 0.048. Abbreviations: $\beta$ = standardized path coefficients, 95% CI = 95% Confidence Interval, SE = Standard Error, $p$ = p-value, PVS = Problematic Video Streaming, CFI = Comparative Fit Index, TLI = Tucker-Lewis Index, RMSEA = Root Mean Square Error of Approximation, SRMR = Standardized Root Mean square Residuals. + $p < .10$ , \* $p < .05$ , \*\* $p < .01$ , \*\*\* $p < .001$ .

## Discussion

This study is the first to provide representative data on the prevalence trends of adolescent PVS in Germany over four consecutive years (2022-2025) using standardized ICD-11 criteria for disorders due to addictive behaviours and distinguishing between hazardous and pathological patterns. Further, this study is the first to explore the temporal relationship between PVS and depression in a longitudinal approach over four years.

### Prevalence of adolescent PVS

The prevalence of adolescent *hazardous VS* was stable at around 13-14% between 2022 and 2024, but almost doubled to 25% in 2025, while estimates for *pathological VS* ranged from 2.6 to 4.2% across waves. These findings are of high clinical and prevention relevance, as the observed prevalence not only exceeds previous estimations [4], but even surpasses global estimates of other PIU [5], contributing to the current discussion and need for further policy responses. The prevalence estimates of *pathological VS* in this study are comparable with national estimates of adolescent *gaming disorder* [27], which previously has been associated with significant impairment in important areas of life for adolescents, necessitating clinical intervention [46,47]. Importantly, the emerging trends of *hazardous VS* may still be amenable to early intervention, offering a critical window to implement targeted preventive strategies.

The marked increase of *hazardous VS* in 2025 and the lack of significant age or sex differences may reflect the growing appeal and availability of video-based entertainment on the internet. Online videos cater to a wide audience through both professionally and user-generated content. Moreover, social media platforms increasingly rely on video-based content, including long-form, short, and live video formats [48], while AI-based personalization algorithms continue to improve [49], and new formats, such as so-called *micro dramas*, are increasingly embraced by audiences [50].

### Association of adolescent PVS and depression

Although *pathological VS* is currently not recognized as a clinical diagnosis, those fulfilling all criteria of a disorder due to addictive behaviours show considerably worse psychological problems compared to healthy controls. According to the results of this study, up to 81% of adolescents with *pathological VS* showed clinically relevant symptoms of depression, compared to 8-9% of non-affected adolescents. Prevalence of clinically relevant symptoms of depression was 2.6 to 3.8 times lower among adolescents with *hazardous VS* (up to 21%), though still more than twice as high compared to non-affected individuals (8-9%). Thus, this study highlights the importance of distinguishing *hazardous* from *pathological VS* and recognizing PVS as a serious public health concern warranting effective prevention and intervention measures. Furthermore, cross-sectional associations between depression and PVS in the CLPA were moderate to large and statistically significant. These findings are consistent with meta-analyses reporting positive, moderate associations between depression and problematic social media use [18], problematic gaming [51], and PIU [52,53] indicating depression to play an important role in the development of PVS.

Consistent with the I-PACE framework, the CLPA conducted in this study suggests depression to significantly predict subsequent PVS in adolescents. However, PVS did not significantly predict depression, which contradicts the argument that (problematic) digital media use may be the preceding factor of the rising mental health issues in youth [13]. Interestingly, the findings are supported by previous research revealing a similar relationship between depression and adolescent PIU [25,54,55].

Depression-related loss of drive commonly results in the inability to engage in pleasant activities. Hence, adolescents suffering from depression may turn to viewing online videos as a low effort opportunity for mood enhancement, comfort, or distraction [56–58]. PVS patterns may then be maintained through addictive design features, such as personalizing recommendations, autoplay, limited availability, and social reinforcements, promoting prolonged engagement [49,59]. Importantly, other research has shown that PIU can predict depression [60] or suggested the relationship to be bidirectional [61,62]. The difference between the present study and previous findings may be due to conceptual differences between PIU and PVS, cultural differences, or influences of exceptional conditions during assessment periods, e.g., the COVID-19 pandemic.

### Clinical implications

Several clinical implications can be drawn, including recognizing PVS as a serious health concern for adolescents and acknowledging depression as an important risk factor. Firstly, mental health professionals who treat adolescents with clinically relevant symptoms of depression should address potential PVS habits due to the elevated rates in this target group and the directional relationship found in this study. Secondly, structured (group) psychotherapy programs that address problematic digital media use may be adapted for adolescents PVS to increase treatment benefits [63,64]. Thirdly, parents should be included in this treatment due to their influence as role-models and facilitators [65]. Lastly, in cases of concurrent occurrence, interventions for depression and PVS should be conducted simultaneously, which is in line with the German guidelines on the treatment of internet use disorders [66–68].

### Strengths & limitations

The robustness of this study stems from its long observation period, the large and representative adolescent samples, and the use of standardized instruments for depression in adolescents [29] and PVS based on criteria derived from ICD-11 criteria for *hazardous gaming* and *gaming disorder* [4]. Another strength of the study is the use of a CLPA within a longitudinal design. This method models autoregressive effects by controlling for prior levels of each variable, allowing the examination of temporal precedence in reciprocal associations. Compared to cross-sectional or simpler longitudinal approaches, this strengthens inferences about the directionality of effects.

Limitations include reliance on self-report measures prone to recall and social desirability bias, and the modest longitudinal sample size, which warranted model simplification. Although random intercept CLPA could not be implemented due to persistent convergence issues, future studies should consider this approach to better capture within-person dynamics [43]. Furthermore, slight asynchrony between waves may have influenced comparability, while the long distances between waves may have influenced prospective associations. Although usual for large population-based surveys, it was not possible to control for potential individual treatment effects. Despite these limitations, the excellent overall model fit strengthens confidence in the present findings. Future research should focus further on examining the role of depression in the emergence of adolescent PVS, utilizing longitudinal designs with larger sample sizes and shorter periods between observations to yield reliable findings on its development and directional relationship over time.

## Conclusion

This study offers timely, representative trend data on adolescent hazardous and pathological online video use (subsumed as PVS) across four consecutive years. By adapting the official ICD-11 criteria for behavioral addictions, it averts overpathologizing a popular digital media activity while pointing out its clinical relevance. Prevalence estimates of *pathological VS* were comparable to those of adolescent *gaming disorder*, while *hazardous VS* estimates even surpassed those of other PIU. Moreover, adolescents with PVS showed markedly higher rates of clinically relevant symptoms of depression across waves – a notable finding within the current debate on the youth mental health crisis. Longitudinally, depression preceded adolescent PVS, but not vice versa. Early public health and policy responses that address depression as a risk factor may prevent further escalation of PVS and its potential long-term consequences. Continued longitudinal research based on established measures is warranted to clarify causal mechanisms between adolescent psychopathology and PVS and inform targeted interventions promoting mental health and digital media literacy.

## Supplementary material

The supplementary material for this article can be found at INSERT LINK.

## Data availability statement

The data supporting the findings of this study are part of an ongoing large study on problematic media use. They are available from the corresponding author upon reasonable request once all results have been published.

## Acknowledgements

The authors would like to thank all study participants and the German *forsa* Institute for Social Research and Statistical Analysis for the data collection.

## Author contribution

JOC and KP developed and conceptualized the overarching study goals, aims, and methods; JOC and KB performed the formal data analyses; JOC was responsible for writing the initial draft of the manuscript; LK, KB, and KP thoroughly reviewed and edited the manuscript draft; LK and KP supervised the manuscript/project; KP acquired funding for and led the project. All authors read, reviewed, and approved the final manuscript.

## Ethics statement

The study was approved by the Local Psychological Ethics Committee at the Center for Psychosocial Medicine of the University Medical Center Hamburg-Eppendorf (UKE) (LPEK-0070) and complied with the Declaration of Helsinki. Written informed consent was obtained from all participants and parents of those aged below 18 years.

## Generative AI statement

During the preparation of this work the authors used *perplexity.ai* in order to improve grammar and wording. After using this service, the authors reviewed and edited the content as needed and take full responsibility for the content of the published article.

## Supplementary Materials

**Figure S1.**
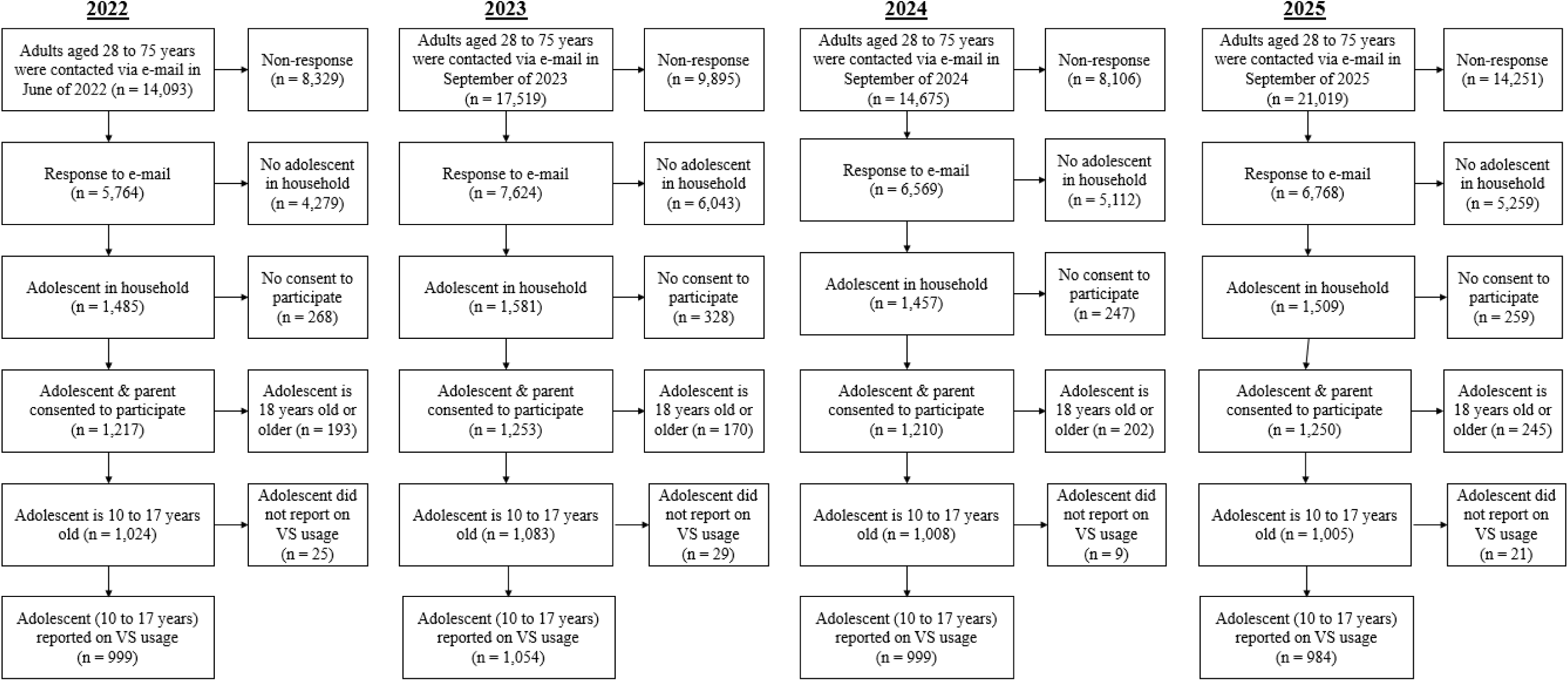
Flow chart of the population-based prevalence estimations.

**Figure S2.**
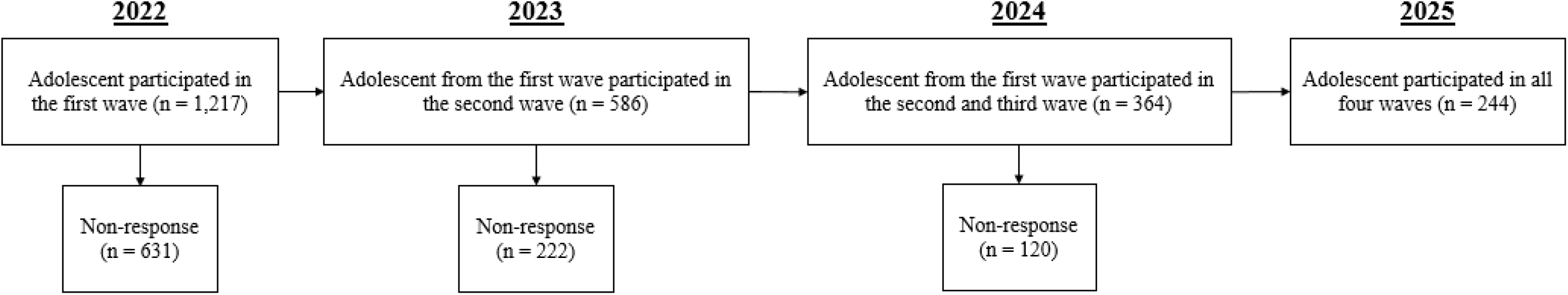
Flow chart of the cross-lagged panel analysis.

**Table S1.** Attrition analysis comparing the baseline sample with the longitudinal sample.

|  | Baseline<br>sample<br>n = 1,217 | Longitudinal<br>sample<br>n = 244 | Sample<br>differences | Difference<br>test | Equivalence<br>test<sup>a</sup> |  |
| --- | --- | --- | --- | --- | --- | --- |
|  | % /<br>M (SD) | % /<br>M (SD) | Mean/<br>Proportion<br>difference | Welch t-test/<br>Difference in two<br>proportions z-test | TOST Lower | TOST Upper |
| **Variables** |  |  |  |  |  |  |
| **Age (in years)** | 14.48 (2.75) | 14.17 (2.65) | 0.31 | t (355.86) = 1.67,<br>*p* = .097<sup>+</sup> | t (355.86) = 3.27,<br>*p* < .001*** | t (355.86) = 0.06,<br>*p* = .523 |
| **Age groups** |  |  |  |  |  |  |
| Early adolescence (10-13 years) | 39.52 | 45.90 | -0.06 | z = 6.78,<br>*p* < .001*** | - | - |
| Mid adolescence (14-17 years) | 44.62 | 40.98 |  |  |  |  |
| Late adolescence (18+ years) | 15.86 | 13.11 |  |  |  |  |
| **Sex** |  |  |  |  |  |  |
| Female | 46.67 | 39.34 | 0.07 | z = -6.59,<br>*p* < .001*** | - | - |
| Male | 53.33 | 60.66 |  |  |  |  |
| **Education** |  |  |  |  |  |  |
| Low | 9.10 | 11.32 | -0.02 | z = 12.69,<br>*p* < .001*** | - | - |
| Medium | 37.09 | 36.79 |  |  |  |  |
| High | 53.81 | 51.89 |  |  |  |  |
| **Place of residence** |  |  |  |  |  |  |
| Urban | 80.84 | 81.97 | -0.01 | z = 10.66,<br>*p* < .001*** | - | - |
| Rural | 19.16 | 18.08 |  |  |  |  |
| **Average video streaming time**<br>**(in hours per week)** | 9.83 (11.99) | 9.14 (9.68) | 0.69 | t (378.72) = 0.94,<br>*p* = .348 | t (378.72) = 1.34,<br>*p* = .090<sup>+</sup> | t (378.72) = 0.54,<br>*p* = .703 |
| **Problematic video-streaming**<br>**(STREDIS-A sum score)** | 5.98 (7.13) | 6.72 (7.42) | -0.74 | t (338.90) = -1.43,<br>*p* = .154 | t (338.90) = -0.85,<br>*p* = .802 | t (338.90) = -2.01,<br>*p* = .023* |
| **Depression**<br>**(PHQ-9 sum score)** | 4.47 (4.70) | 4.36 (4.42) | 0.11 | t (361.89) = 0.36,<br>*p* = .718 | t (361.89) = 1.32,<br>*p* = .094 | t (361.89) = -0.60,<br>*p* = .276 |

*Note.* Longitudinal sample was compared to baseline sample (W1). <sup>a</sup>Bounds were set at $\Delta = \pm 0.3$ for 90% power and $\alpha = 0.05$ . Abbreviations: n = sample size; % = relative frequency; M = arithmetic mean; SD = Standard Deviation, TOST = Two One-Sided Tests, STREDIS-A = Streaming Disorder Scale for Adolescents; PHQ-9 = Patient Health Questionnaire 9-item version, + *p* < .10, \* *p* < .05, \*\* *p* < .01, \*\*\* *p* < .001.

**Table S2.** Sociographic characteristics of German adolescents [representative data, non-imputed].

|  | **2022** | **2023** | **2024** | **2025** |
| --- | --- | --- | --- | --- |
|  | n = 999 | n = 1,054 | n = 999 | n = 984 |
|  | % / | % / | % / | % / |
| **Variables** | M (SD) | M (SD) | M (SD) | M (SD) |
| **Age** (in years) | 13.5 (2.2) | 13.6 (2.3) | 13.5 (2.3) | 13.5 (2.2) |
| Young adolescents (10-13 years) | 49.93 | 49.45 | 50.08 | 49.72 |
| Older adolescents (14-17 years) | 50.07 | 50.55 | 49.92 | 50.28 |
| **Sex** |  |  |  |  |
| Female | 48.39 | 48.64 | 48.55 | 50.30 |
| Male | 51.61 | 51.36 | 51.45 | 49.70 |
| **Education** |  |  |  |  |
| Low | 11.28 | 13.08 | 11.28 | 7.95 |
| Medium | 38.81 | 30.17 | 32.08 | 32.74 |
| High | 49.91 | 56.75 | 56.63 | 59.31 |
| NA | 69 | 91 | 87 | 116 |
| **Place of residence** |  |  |  |  |
| Urban | 79.94 | 79.27 | 79.81 | 78.79 |
| Rural | 20.06 | 20.73 | 20.19 | 21.21 |
| NA | 1 | 0 | 0 | 0 |
| **Average video streaming time** (in hours per week)<sup>a</sup> | 8.8 (11.1) | 9.6 (11.5) | 9.4 (12.9) | 10.9 (13.0) |
| NA | 92 | 83 | 140 | 87 |
| **Problematic video streaming** (STREDIS-A sum score) | 6.3 (7.7) | 6.7 (7.2) | 7.0 (7.4) | 8.2 (7.3) |
| NA | 20 | 25 | 36 | 26 |
| **Depression** (PHQ-9 sum score) | 4.5 (5.1) | 4.8 (4.9) | 4.9 (4.6) | 4.7 (5.0) |
| NA | 56 | 73 | 55 | 49 |

*Note.* Based on weighted, non-imputed data. <sup>a</sup>Based on regular users only (i.e., reported to watch videos at least once per week). Abbreviations: n = sample size, % = relative frequency, M = arithmetic mean, SD = Standard Deviation, NA = missing data, STREDIS-A = Streaming Disorder Scale for Adolescents, PHQ-9 = Patient Health Questionnaire 9-item version.

**Table S3.** Sensitivity analyses (complete-case analysis) using non-imputed data from PVS prevalence estimates.

| Groups | 2022<br>n = 999 |  |  |  | 2023<br>n = 1,054 |  |  |  |
| --- | --- | --- | --- | --- | --- | --- | --- | --- |
|  | Estimate %<br>[95 % CI] |  |  |  | Estimate %<br>[95 % CI] |  |  |  |
|  | Hazardous<br>VS<br>non-imputed<br>+ weighted | Hazardous<br>VS<br>imputed +<br>weighted | Pathological<br>VS<br>non-imputed<br>+ weighted | Pathological<br>VS<br>imputed +<br>weighted | Hazardous<br>VS<br>non-imputed<br>+ weighted | Hazardous<br>VS<br>imputed +<br>weighted | Pathological<br>VS<br>non-imputed<br>+ weighted | Pathological<br>VS<br>imputed +<br>weighted |
| **Total** | 14.36<br>[12.25; 16.65] | 14.41<br>[12.21; 16.61] | 2.56<br>[1.69; 3.67] | 2.62<br>[1.63; 3.62] | 12.71<br>[10.76; 14.85] | 12.82<br>[10.77; 14.86] | 4.10<br>[3.00; 5.344] | 4.24<br>[3.01; 5.46] |
| **Sex:** |  |  |  |  |  |  |  |  |
| Female | 16.59<br>[13.50; 20.23] | 16.54<br>[13.19; 19.89] | 3.16<br>[1.91; 5.19] | 3.10<br>[1.55; 4.65] | 15.30<br>[12.40; 18.74] | 15.35<br>[12.18; 18.53] | 4.37<br>[2.88; 6.56] | 4.70<br>[2.85; 6.55] |
| Male | 12.27<br>[9.68; 15.43] | 12.41<br>[9.55; 15.28] | 1.99<br>[1.08; 3.66] | 2.17<br>[0.91; 3.44] | 10.27<br>[7.95; 13.17] | 10.42<br>[7.79; 13.04] | 3.86<br>[2.51; 5.88] | 3.80<br>[2.18; 5.41] |
| **Age:** |  |  |  |  |  |  |  |  |
| 10-13 years | 14.23<br>[11.42; 17.61] | 14.23<br>[11.15; 17.32] | 2.26<br>[1.26; 4.02] | 2.48<br>[1.11; 3.84] | 14.23<br>[11.44; 17.55] | 14.48<br>[11.39; 17.57] | 3.20<br>[1.97; 5.14] | 3.22<br>[1.69; 4.76] |
| 14-17 years | 14.48<br>[11.61; 17.91] | 14.59<br>[11.45; 17.73] | 2.86<br>[1.69; 4.78] | 2.77<br>[1.33; 4.21] | 11.23<br>[8.78; 14.25] | 11.19<br>[8.46; 13.92] | 4.99<br>[3.41; 7.24] | 5.23<br>[3.33; 7.14] |
| **Depression:** |  |  |  |  |  |  |  |  |
| No (PHQ-9 < 10) | 13.16<br>[11.01; 15.66] | 13.11<br>[10.83; 15.39] | 0.55<br>[0.22; 1.37] | 0.58<br>[0.07; 1.08] | 11.28<br>[9.31; 13.60] | 11.70<br>[9.58; 13.82] | 1.14<br>[0.61; 2.13] | 1.23<br>[0.51; 1.94] |
| Yes (PHQ-9 ≥ 10) | 24.40<br>[17.39; 33.11] | 23.01<br>[15.72; 30.31] | 15.94<br>[10.30; 23.85] | 16.13<br>[9.81; 22.44] | 20.59<br>[14.29; 28.75] | 20.09<br>[13.02; 27.16] | 23.26<br>[16.57; 31.64] | 23.84<br>[16.69; 30.99] |

*Note.* Non-imputed data refers to observed data only (i.e., complete-case analysis). Imputed + weighted refers to weighted prevalence estimates based on pooled results across multiple imputed datasets (m = 10). Abbreviations: PVS = Problematic Video Streaming, 95% CI = 95% Confidence Interval, VS = Video Streaming, n = sample size, PHQ-9 = Patient Health Questionnaire 9-item version.

**Table S3.** Sensitivity analyses (complete-case analysis) using non-imputed data from PVS prevalence estimates (continued).
| Groups | 2024<br>n = 999 |  |  |  | 2025<br>n = 984 |  |  |  |
| --- | --- | --- | --- | --- | --- | --- | --- | --- |
|  | Estimate %<br>[95 % CI] |  |  |  | Estimate %<br>[95 % CI] |  |  |  |
|  | Hazardous<br>VS<br>non-imputed<br>+ weighted | Hazardous<br>VS<br>imputed +<br>weighted | Pathological<br>VS<br>non-imputed<br>+ weighted | Pathological<br>VS<br>imputed +<br>weighted | Hazardous<br>VS<br>non-imputed<br>+ weighted | Hazardous<br>VS<br>imputed +<br>weighted | Pathological<br>VS<br>non-imputed<br>+ weighted | Pathological<br>VS<br>imputed +<br>weighted |
| **Total** | 14.00<br>[11.90; 16.30] | 14.21<br>[11.97; 16.44] | 3.24<br>[2.24; 4.49] | 3.33<br>[2.20; 4.47] | 24.79<br>[22.13; 27.59] | 25.11<br>[22.39; 27.82] | 3.71<br>[2.64; 5.04] | 3.80<br>[2.59; 5.01] |
| **Sex:** |  |  |  |  |  |  |  |  |
| Female | 12.88<br>[10.12; 16.25] | 13.18<br>[10.00; 16.36] | 4.39<br>[2.86; 6.69] | 4.21<br>[2.42; 6.01] | 26.46<br>[22.73; 30.56] | 26.76<br>[22.84; 30.67] | 2.68<br>[1.56; 4.55] | 2.63<br>[1.22; 4.03] |
| Male | 15.05<br>[12.16; 18.49] | 15.18<br>[12.02; 18.33] | 2.15<br>[1.16; 3.88] | 2.51<br>[1.10; 3.91] | 23.07<br>[19.59; 27.09] | 23.44<br>[19.67; 27.21] | 4.78<br>[3.19; 7.12] | 4.99<br>[3.02; 6.97] |
| **Age:** |  |  |  |  |  |  |  |  |
| 10-13 years | 14.88<br>[11.97; 18.35] | 14.75<br>[11.59; 17.92] | 2.26<br>[1.26; 4.05] | 2.53<br>[1.13; 3.94] | 26.70<br>[22.91; 30.88] | 27.36<br>[23.40; 31.33] | 3.32<br>[2.03; 5.37] | 3.27<br>[1.68; 4.86] |
| 14-17 years | 13.11<br>[10.35; 16.46] | 13.66<br>[10.48; 16.84] | 4.22<br>[2.74; 6.45] | 4.14<br>[2.37; 5.90] | 22.93<br>[19.41; 26.88] | 22.88<br>[19.17; 26.58] | 4.10<br>[2.66; 6.28] | 4.33<br>[2.51; 6.15] |
| **Depression:** |  |  |  |  |  |  |  |  |
| No (PHQ-9 < 10) | 12.47<br>[10.38; 14.92] | 12.86<br>[10.61; 15.11] | 1.76<br>[1.06; 2.93] | 1.79<br>[0.91; 2.67] | 23.31<br>[20.51; 26.36] | 23.63<br>[20.75; 26.51] | 1.37<br>[0.76; 2.45] | 1.37<br>[0.58; 2.17] |
| Yes (PHQ-9 ≥ 10) | 22.78<br>[15.42; 32.33] | 24.56<br>[16.25; 32.86] | 16.25<br>[10.09; 25.13] | 15.24<br>[8.41; 22.06] | 31.89<br>[23.97; 41.00] | 34.73<br>[26.15; 43.31] | 21.43<br>[14.83; 29.94] | 19.61<br>[12.66; 26.56] |

*Note.* Non-imputed data refers to observed data only (i.e., complete-case analysis). Imputed + weighted refers to weighted prevalence estimates based on pooled results across multiple imputed datasets ( $m = 10$ ). Abbreviations: PVS = Problematic Video Streaming, 95% CI = 95% Confidence Interval, VS = Video Streaming, $n$ = sample size, PHQ-9 = Patient Health Questionnaire 9-item version.

## References

[1] Statista. Online Video & Entertainment 2025. https://www.statista.com/mar-kets/424/topic/542/online-video-entertainment/#overview.

[2] Casey BJ, Jones RM. Neurobiology of the Adolescent Brain and Behavior: Implications for Substance Use Disorders. J Am Acad Child Adolesc Psychiatry 2010;49:1189–201. 10.1016/j.jaac.2010.08.017.

[3] Yuan K, Qin W, Yu D, Bi Y, Xing L, Jin C, et al. Core brain networks interactions and cognitive control in internet gaming disorder individuals in late adolescence/early adulthood. Brain Struct Funct 2016;221:1427–42. 10.1007/s00429-014-0982-7.

[4] Paschke K, Napp A-K, Thomasius R. Applying ICD-11 criteria of Gaming Disorder to identify problematic video streaming in adolescents: Conceptualization of a new clinical phenomenon. J Behav Addict 2022;11:451–66. 10.1556/2006.2022.00041.

[5] Boniel-Nissim M, Marino C, Galeotti T, Blinka L, Ozoliņa K, Craig W, et al. A focus on adolescent social media use and gaming in Europe, central Asia and Canada. Health Behaviour in School-aged Children international report from the 2021/2022 survey. 2024. https://www.who.int/europe/news/item/25-09-2024-teens--screens-and-mental-health.

[6] American Psychological Association. APA Recommendations for Healthy Teen Video Viewing. A Summary of the Science with Action Steps regarding Video Viewing and Adolescent Well-Being 2024. https://www.apa.org/topics/social-media-inter-net/healthy-teen-video-viewing.

[7] Rahat M, Mojgani J, Lethbridge G, Al-Bya H, Patterson B, Goldman Bergmann C, et al. Problematic video-streaming: a short review. Curr Opin Behav Sci 2022;48:101232. 10.1016/j.cobeha.2022.101232.

[8] Paulus FW, Joas J, Gerstner I, Kühn A, Wenning M, Gehrke T, et al. Problematic Internet Use among Adolescents 18 Months after the Onset of the COVID-19 Pandemic. Children 2022;9:1724. 10.3390/children9111724.

[9] Fineberg NA, Potenza MN. Addressing problematic use of the Internet and related compulsive and addictive behaviors. Curr Opin Behav Sci 2023;51:101279. 10.1016/j.cobeha.2023.101279.

[10] World Health Organization. Clinical descriptions and diagnostic requirements for ICD-11 mental, behavioural and neurodevelopmental disorders (CDDR). Geneva, Switzerland: World Health Organization; 2024.

[11] Brand M, Rumpf H-Jü, Demetrovics Z, Müller A, Stark R, King DL, et al. Which conditions should be considered as disorders in the International Classification of Diseases (ICD-11) designation of “other specified disorders due to addictive behaviors”? J Behav Addict 2020;11:150–9. 10.1556/2006.2020.00035.

[12] Paschke K, Napp A-K, Thomasius R. Parents Rate Problematic Video Streaming in Adolescents: Conceptualization and External Assessment of a New Clinical Phenomenon Based on the ICD-11 Criteria of Gaming Disorder. J Clin Med 2023;12:1010. 10.3390/jcm12031010.

[13] McGorry P, Gunasiri H, Mei C, Rice S, Gao CX. The youth mental health crisis: analysis and solutions. Front Psychiatry 2025;15:1517533. 10.3389/fpsyt.2024.1517533.

[14] Alimoradi Z, Jafari E, Potenza MN, Lin C-Y, Wu C-Y, Pakpour AH. Binge-Watching and Mental Health Problems: A Systematic Review and Meta-Analysis. Int J Environ Res Public Health 2022;19:9707. 10.3390/ijerph19159707.

[15] Starosta JA, Izydorczyk B. Understanding the Phenomenon of Binge-Watching—A Systematic Review. Int J Environ Res Public Health 2020;17:4469. 10.3390/ijerph17124469.

[16] Chao M, Lei J, He R, Jiang Y, Yang H. TikTok use and psychosocial factors among adolescents: Comparisons of non-users, moderate users, and addictive users. Psychiatry Res 2023;325:115247. 10.1016/j.psychres.2023.115247.

[17] Keles B, McCrae N, Grealish A. A systematic review: the influence of social media on depression, anxiety and psychological distress in adolescents. Int J Adolesc Youth 2020;25:79–93. 10.1080/02673843.2019.1590851.

[18] Shannon H, Bush K, Villeneuve PJ, Hellemans KG, Guimond S. Problematic Social Media Use in Adolescents and Young Adults: Systematic Review and Meta-analysis. JMIR Ment Health 2022;9:e33450. 10.2196/33450.

[19] Sugaya N, Shirasaka T, Takahashi K, Kanda H. Bio-psychosocial factors of children and adolescents with internet gaming disorder: a systematic review. Biopsychosoc Med 2019;13:3. 10.1186/s13030-019-0144-5.

[20] Cairns KE, Yap MBH, Pilkington PD, Jorm AF. Risk and protective factors for depression that adolescents can modify: A systematic review and meta-analysis of longitudinal studies. J Affect Disord 2014;169:61–75. 10.1016/j.jad.2014.08.006.

[21] Kassis W, Artz S, White J. Understanding Depression in Adolescents: A Dynamic Psychosocial Web of Risk and Protective Factors. Child Youth Care Forum 2017;46:721–43. 10.1007/s10566-017-9404-3.

[22] Brand M, Wegmann E, Stark R, Müller A, Wölfling K, Robbins TW, et al. The Interaction of Person-Affect-Cognition-Execution (I-PACE) model for addictive behaviors: Update, generalization to addictive behaviors beyond internet-use disorders, and specification of the process character of addictive behaviors. Neurosci Biobehav Rev 2019;104:1–10. 10.1016/j.neubiorev.2019.06.032.

[23] Brand M, Antons S, Bőthe B, Demetrovics Z, Fineberg NA, Jimenez-Murcia S, et al. Current Advances in Behavioral Addictions: From Fundamental Research to Clinical Practice. Am J Psychiatry 2025;182:155–63. 10.1176/appi.ajp.20240092.

[24] Coutelle R, Balzer J, Rolling J, Lalanne L. Problematic gaming, psychiatric comorbid-ities, and adolescence: A systematic review of the literature. Addict Behav 2024;157:108091. 10.1016/j.addbeh.2024.108091.

[25] Leo K, Kewitz S, Wartberg L, Lindenberg K. Depression and Social Anxiety Predict Internet Use Disorder Symptoms in Children and Adolescents at 12-Month Follow-Up: Results From a Longitudinal Study. Front Psychol 2021;12:787162. 10.3389/fpsyg.2021.787162.

[26] Pazdur M, Tutus D, Haag A-C. Risk Factors for Problematic Social Media Use in Youth: A Systematic Review of Longitudinal Studies. Adolesc Res Rev 2025;10:237–53. 10.1007/s40894-025-00264-4.

[27] Busch K, Wiedemann H, Klamert L, Paschke K. Epidemiology of Adolescent Gaming Disorder: A Six-Year Population-Based Longitudinal Study (2019-2024). Eur Psychiatry 2026:1–28. 10.1192/j.eurpsy.2026.12232.

[28] Wiedemann H, Lüdecke D, Thomasius R, Paschke K. Six-year trends in problematic social media use prevalence and sociodemographic predictors in German adolescents (2019–2024). Comput Hum Behav Rep 2026;21:100958. 10.1016/j.chbr.2026.100958.

[29] Kroenke K, Spitzer RL, Williams JBW. The PHQ-9: Validity of a brief depression severity measure. J Gen Intern Med 2001;16:606–13. 10.1046/j.1525-1497.2001.016009606.x.

[30] Manea L, Gilbody S, McMillan D. Optimal cut-off score for diagnosing depression with the Patient Health Questionnaire (PHQ-9): a meta-analysis. Can Med Assoc J 2012;184:E191–6. 10.1503/cmaj.110829.

[31] Sawyer SM, Azzopardi PS, Wickremarathne D, Patton GC. The age of adolescence. Lancet Child Adolesc Health 2018;2:223–8. 10.1016/S2352-4642(18)30022-1.

[32] Deng Y, Li Y, Chen H, Li M, Tao Y. A network approach to personality vulnerability symptom structure across early, middle, and late adolescence: Insights from a large-scale sample. J Affect Disord 2025;378:155–64. 10.1016/j.jad.2025.02.087.

[33] UNESCO Institute for Statistics. International standard classification of education: ISCED 2011 2012. https://uis.unesco.org/sites/default/files/documents/international-standard-classification-of-education-isced-2011-en.pdf. (accessed July 17, 2026).

[34] BBSR. Stadt- und Gemeindetypen in Deutschland. Bundesinstitut für Bau-, Stadt- und Raumforschung. 2020. https://www.bbsr.bund.de/BBSR/DE/themen/stadt-region/staedtesystem-stadttypen/_node.html (accessed July 17, 2026).

[35] R Core Team. R: A Language and Environment for Statistical Computing. Vienna, Austria: R Foundation for Statistical Computing; 2019.

[36] Kim H-Y. Statistical notes for clinical researchers: assessing normal distribution (2) using skewness and kurtosis. Restor Dent Endod 2013;38:52–4. 10.5395/rde.2013.38.1.52.

[37] Revelle W. psych: Procedures for Psychological, Psychometric, and Personality Research 2025.

[38] Buuren S, Groothuis-Oudshoorn K. mice: Multivariate Imputation by Chained Equations in R. J Stat Softw 2011;45:1–67. 10.18637/jss.v045.i03.

[39] Rubin DB. Multiple Imputation for Nonresponse in Surveys. 1st ed. Wiley; 1987. 10.1002/9780470316696.

[40] Makowski D, Ben-Shachar MS, Wiernik BM, Patil I, Thériault R, Lüdecke D. modelbased: An R package to make the most out of your statistical models through marginal means, marginal effects, and model predictions. J Open Source Softw 2025;10:7969. 10.21105/joss.07969.

[41] Statistisches Bundesamt. Fortschreibung des Bevölkerungsstandes 2022. https://www.destatis.de/DE/Themen/Gesellschaft-Umwelt/Bevoelkerung/Bevoelke-rungsstand/_inhalt.html (accessed July 17, 2026).

[42] Rosseel Y, Jorgensen TD, De Wilde L. lavaan: Latent Variable Analysis 2012:0.6-20. 10.32614/CRAN.package.lavaan.

[43] Hamaker EL, Kuiper RM, Grasman RPPP. A critique of the cross-lagged panel model. Psychol Methods 2015;20:102–16. 10.1037/a0038889.

[44] Hu L, Bentler PM. Cutoff criteria for fit indexes in covariance structure analysis: Conventional criteria versus new alternatives. Struct Equ Model Multidiscip J 1999;6:1–55. 10.1080/10705519909540118.

[45] Orth U, Meier LL, Bühler JL, Dapp LC, Krauss S, Messerli D, et al. Effect size guidelines for cross-lagged effects. Psychol Methods 2024;29:421–33. 10.1037/met0000499.

[46] Paschke K, Austermann MI, Thomasius R. Assessing ICD-11 Gaming Disorder in Adolescent Gamers: Development and Validation of the Gaming Disorder Scale for Adolescents (GADIS-A). J Clin Med 2020;9:993. 10.3390/jcm9040993.

[47] Stevens MW, Dorstyn D, Delfabbro PH, King DL. Global prevalence of gaming disorder: A systematic review and meta-analysis. Aust N Z J Psychiatry 2021;55:553–68. 10.1177/0004867420962851.

[48] Statista. Media usage among teenagers in Germany - statistics & facts 2025. https://www.statista.com/topics/5065/media-usage-among-teenagers-in-germany/#top-icOverview.

[49] Flayelle M, Brevers D, King DL, Maurage P, Perales JC, Billieux J. A taxonomy of technology design features that promote potentially addictive online behaviours. Nat Rev Psychol 2023;2:136–50. 10.1038/s44159-023-00153-4.

[50] Rossmanith B. Neue Formate - geteilte Aufmerksamkeit? Micro-Dramen und Long-form-Serien 2026. https://www.youtube.com/watch?v=K211tzalI6E.

[51] Zheng C, Liu K, Liang L, Yuan T, Li H, Qing Y, et al. Revisiting the Relationship Between Adolescent Internet Gaming Disorder and Depression: A Systematic Review and Meta-Analysis. Adolesc Res Rev 2025. 10.1007/s40894-025-00275-1.

[52] Cai Z, Mao P, Wang Z, Wang D, He J, Fan X. Associations Between Problematic Internet Use and Mental Health Outcomes of Students: A Meta-analytic Review. Adolesc Res Rev 2023;8:45–62. 10.1007/s40894-022-00201-9.

[53] Lozano-Blasco R, Cortés-Pascual A. Problematic Internet uses and depression in adolescents: A meta-analysis. Comunicar 2020;28:109–20. 10.3916/C63-2020-10.

[54] Falcione K, Weber R. Psychopathology and Gaming Disorder in Adolescents. JAMA Netw Open 2025;8:e2528532. 10.1001/jamanetworkopen.2025.28532.

[55] Raudsepp L, Kais K. Longitudinal associations between problematic social media use and depressive symptoms in adolescent girls. Prev Med Rep 2019;15:100925. 10.1016/j.pmedr.2019.100925.

[56] Kircaburun K, Harris A, Calado F, Griffiths MD. Emotion regulation difficulties and problematic mukbang watching: The mediating role of psychological distress and impulsivity. Psychiatry Res Commun 2024;4:100152. 10.1016/j.psy-com.2023.100152.

[57] Myrick JG. Emotion regulation, procrastination, and watching cat videos online: Who watches Internet cats, why, and to what effect? Comput Hum Behav 2015;52:168–76. 10.1016/j.chb.2015.06.001.

[58] Starosta J, Izydorczyk B, Sitnik-Warchulska K, Lizińczyk S. Impulsivity and Difficulties in Emotional Regulation as Predictors of Binge-Watching Behaviours. Front Psychiatry 2021;12:743870. 10.3389/fpsyt.2021.743870.

[59] Chaudhary A, Saroha J, Monteiro K, Forbes AG, Parnami A. “Are You Still Watching?”: Exploring Unintended User Behaviors and Dark Patterns on Video Streaming Platforms. Des. Interact. Syst. Conf., Virtual Event Australia: ACM; 2022, p. 776–91. 10.1145/3532106.3533562.

[60] Kojima R, Shinohara R, Akiyama Y, Yokomichi H, Yamagata Z. Temporal directional relationship between problematic internet use and depressive symptoms among Japanese adolescents: A random intercept, cross-lagged panel model. Addict Behav 2021;120:106989. 10.1016/j.addbeh.2021.106989.

[61] Fang X, Tian M, Wang R, Wang P. Relationships between depression, loneliness and pathological Internet use in adolescents: A cross-lagged analysis. Curr Psychol 2023;42:20696–706. 10.1007/s12144-022-03180-1.

[62] Morita M, Ando S, Kiyono T, Morishima R, Yagi T, Kanata S, et al. Bidirectional relationship of problematic Internet use with hyperactivity/inattention and depressive symptoms in adolescents: a population-based cohort study. Eur Child Adolesc Psychiatry 2022;31:1601–9. 10.1007/s00787-021-01808-4.

[63] Cloes J-O, Bitter K, Lorenz L, Thomasius R, Paschke K. Resource-strengthening training for adolescents with problematic gaming (Res@t-A): Results from an exploratory single-arm pre-post-follow-up intervention study. J Behav Addict 2026:2006.2025.00211. 10.1556/2006.2025.00211.

[64] Paschke K, Diestelkamp S, Zapf A, Busch K, Arnaud N, Prehn-Kristensen A, et al. An app-based training for adolescents with problematic digital-media use and their parents (Res@t digital): protocol for a cluster-randomized clinical trial. Front Psychiatry 2024;14:1245536. 10.3389/fpsyt.2023.1245536.

[65] Hülquist J, Fangerau N, Thomasius R, Paschke K. Resource-Strengthening Training for Parents of Adolescents with Problematic Gaming (Res@t-P): A Clinical Pilot Study. Int J Environ Res Public Health 2022;19:9495. 10.3390/ijerph19159495.

[66] Müller KW, Bilke-Hentsch O, Bottel L, Noack M, Scherer L, Schneider K, et al. Guidelines on the Treatment of Social Networking Sites Use Disorder. SUCHT 2025;71:117–22. 10.1024/0939-5911/a000920.

[67] Wildt BT, Müller KW, Dieris-Hirche J, Feindel H, Lindenberg K, Noack M, et al. Guidelines on the Treatment of Gaming Disorder. SUCHT 2025;71:93–101. 10.1024/0939-5911/a000922.

[68] Wölfling K, Eichenberg C, Leménager T, Basenach L, Dreier M, Rüther T, et al. Guideline on the Treatment of General Internet Use Disorder. SUCHT 2025;71:87–92. 10.1024/0939-5911/a000918.

